# Histo-blood Group Antigen status of Australian Aboriginal children and seropositivity following oral rotavirus vaccination

**DOI:** 10.1101/2022.11.24.22282699

**Authors:** Bianca F. Middleton, Margie Danchin, Nigel A. Cunliffe, Mark A. Jones, Karen Boniface, Carl D. Kirkwood, Sarah Gallagher, Lea-Ann Kirkham, Caitlyn Granland, Monica McNeal, Celeste Donato, Nada Bogdanovic-Sakran, Amanda Handley, Julie E. Bines, Thomas L. Snelling

**Affiliations:** Global and Tropical Health Division, Menzies School of Health Research, Charles Darwin University, Darwin, Australia; Vaccine Uptake Group, Murdoch Children’s Research Institute, Melbourne, Australia; Department of Paediatrics, University of Melbourne, Melbourne, Australia; Department of General Medicine, Royal Children’s Hospital, Melbourne, Australia; Department of Clinical Infection, Microbiology and Immunology, University of Liverpool, Liverpool, UK; Health and Clinical Analytics, School of Public Health, University of Sydney, Sydney, Australia; Infection and Immunity, Murdoch Children’s Research Institute, Melbourne, Australia; Enteric and Diarrheal Diseases, Bill and Melinda Gates Foundation, Seattle, USA; Wesfarmers Centre of Vaccines and Infectious Diseases, Telethon Kids Institute, Perth, Australia; Centre for Child Health Research, University of Western Australia, Perth, Australia; Department of Pediatrics, University of Cincinnati College of Medicine, Cincinnati, USA; Division of Infectious Disease, Cincinnati Children’s Hospital Medical Centre, Cincinnati, USA; Department of Gastroenterology, The Royal Children’s Hospital, Melbourne, Victoria, Australia; School of Public Health, Curtin University, Perth, Australia

## Abstract

**Background:** High rates of breakthrough rotavirus gastroenteritis have been reported among Aboriginal children living in rural and remote Australia despite receipt of two doses of oral rotavirus vaccine. Histo-blood group antigens (HBGAs) may mediate rotavirus genotype-dependent differences in susceptibility to rotavirus infection and immune responses to rotavirus vaccination.

**Methods:** HBGA phenotype – Lewis and secretor status - was determined by enzyme immunoassay of saliva samples obtained from Australian Aboriginal children who were enrolled at age 6 to <12 months in a randomised clinical trial of an additional (booster) dose of oral rotavirus vaccine. Participants had received the routine two-dose schedule of oral rotavirus vaccine administered at age 6 weeks and 4 months. Non-secretor phenotype was confirmed by DNA extraction to identify *FUT2* ‘G428A’ mutation. Rotavirus seropositivity was defined as serum anti-rotavirus IgA ≥ 20 AU/mL measured by ELISA on enrolment.

**Results:** Of 156 children, 119 (76%) were secretors, 129 (83%) were Lewis antigen positive, and 105 (67%) were rotavirus IgA seropositive. Eighty-seven of 119 (73%) secretors were rotavirus seropositive, versus 4/9 (44%) weak secretors and 13/27 (48%) non-secretors. Eighty-nine of 129 (69%) Lewis antigen positive children were rotavirus seropositive versus 10 of 19 (53%) of those who were Lewis antigen negative.

**Conclusions:** Most Australian Aboriginal children were secretor and Lewis antigen positive. Non-secretor children were less likely to be seropositive for rotavirus following vaccination, but this phenotype was less common. HBGA status is unlikely to fully explain the underperformance of rotavirus vaccine at a population level among Australian Aboriginal children.

## Introduction

Rotavirus remains a leading cause of dehydrating diarrhoeal disease in young children. The introduction of oral rotavirus vaccines into childhood immunization programmes has resulted in a significant reduction in the global burden of rotavirus disease, however vaccine effectiveness has varied across settings. A systematic review of the performance of four oral rotavirus vaccines in high child mortality settings in Asia and Africa reported that vaccine efficacy was 48 to 57% in the first year following vaccination, and 29% to 54% in the second year.^1^ Reduced protection from oral rotavirus vaccines has also been reported among rural and remote Australian Aboriginal and Torres Strait Islander children.^2^ The suboptimal protection from oral rotavirus vaccines observed in these settings has been variously attributed to high levels of maternally derived vaccine-neutralising anti-rotavirus antibodies transferred to infants via the placenta and breast-milk, poor infant nutrition, environmental enteropathy and intestinal dysbiosis, comorbid infection, and a high diversity of circulating rotavirus strains.^3^ Histo-blood group antigens (HBGAs) appear to be an important determinant of host-susceptibility to rotavirus infection and may affect the immune response to oral rotavirus vaccines.^4^

HBGAs are carbohydrates expressed on the surface of gut epithelium and in soluble forms in saliva and breast milk. Synthesis of secretor and Lewis HBGAs is catalysed by glycosyltransferase enzymes encoded by the *FUT2* and *FUT3* genes, respectively. Mutations in these genes can give rise to non-functional enzymes, and subsequently non-secretor and Lewis negative phenotypes. Differences in the prevalence of polymorphisms in the *FUT2* and *FUT3* genes gives rise to differential expression of secretor and Lewis status between populations.^5^

The increasing evidence for the role that HBGA phenotype plays in susceptibility to rotavirus infection has stimulated interest in the possible role that HBGA may play in the immune response to oral rotavirus vaccines. All licensed rotavirus vaccines comprise live attenuated virus strains so it is plausible that factors which affect wild-type rotavirus infection may also apply to rotavirus vaccine strains, as vaccine virus replication within intestinal epithelial cells is required to induce local gut immunity and subsequent vaccine effects.^6^ For the first time, this study reports the relationship between the HBGA status of Australian Aboriginal children and their seropositivity to rotavirus following receipt of a standard two-dose schedule of oral Rotarix rotavirus vaccine.

## Methods

### Study Design & Setting

The study was nested in ORVAC, a double-blind randomised, placebo-controlled clinical trial evaluating the immunological and clinical effect of administering an additional (booster) dose of oral Rotarix rotavirus vaccine to Australian Aboriginal and Torres Strait Islander children aged 6 to < 12 months old. The ORVAC protocol has been published elsewhere.^7^ In brief, children were eligible for enrolment if they were 6 to < 12 months old, identified as Aboriginal and/or Torres Strait Islander, and had been previously been vaccinated with at least one dose of oral Rotarix vaccine (scheduled at age 6 weeks and 4 months old). In ORVAC Stage 1, serum was collected at enrolment to determine IgA seropositivity to rotavirus (at age 6 to < 12 months old) and consent was also sought from a subset of participants to collect a sample of the child’s saliva at enrolment. Approvals were obtained from the Northern Territory Department of Health and Menzies School of Health Research Ethics Committee (2020-3759). The ORVAC protocol is registered on ClinicalTrials.gov (NCT02941107).

### Blood Samples

At enrolment, a blood sample of 1.2 mL was collected to measure anti-rotavirus serum IgA levels. As detailed previously,^7^ specific rotavirus IgA antibodies were measured by enzyme linked immuno-assay using rabbit anti-rotavirus polyclonal antisera as the coating antibody to capture a rotavirus lysate (G1P8) strain. Concentrations of rotavirus-specific IgA were measured in patient serum samples using a reference standard having been assigned a concentration of 1000 arbitrary units (AU/mL). Seropositivity was defined as anti-rotavirus IgA ≥ 20 AU/mL.

### Saliva Samples

At enrolment, 1 mL saliva sample was collected in the GeneFixSaliva DNA Collection tube (Isolhelix, GFXA-01) containing 1 mL of stabilisation buffer. The samples were mixed then divided into two. Aliquot One (1 mL Saliva/Buffer) was used for HBGA phenotyping by enzyme immunoassay - secretor and Lewis antigen status.^8^ Aliquot Two (1 mL Saliva/Buffer) was used for DNA extraction from samples with either non-secretor or indeterminate phenotype results, with PCR and restriction fragment length polymorphism used to identify the FUT2 ‘G428A’ mutation.^8^

### Statistics

Frequencies of anti-rotavirus IgA seropositivity at enrolment by Lewis, secretor and combined Lewis and secretor status were expressed as proportions and percentages, with the average concentration expressed as the geometric mean concentration (GMC). A non-informative beta conjugate prior to the binomial likelihood was used to estimate posterior distributions by phenotype classification and to derive relative risk ratios.

## Results

There were 253 children enrolled into ORVAC Stage 1 between March 2018 and August 2020. Of these, 156 children had saliva collected for HBGA analysis. Baseline demographic characteristics were comparable to the total ORVAC Stage 1 cohort (Appendix 1) and almost all children (96%) had received a complete two-dose course of oral Rotarix vaccine (Table 1).

**Table 1:**
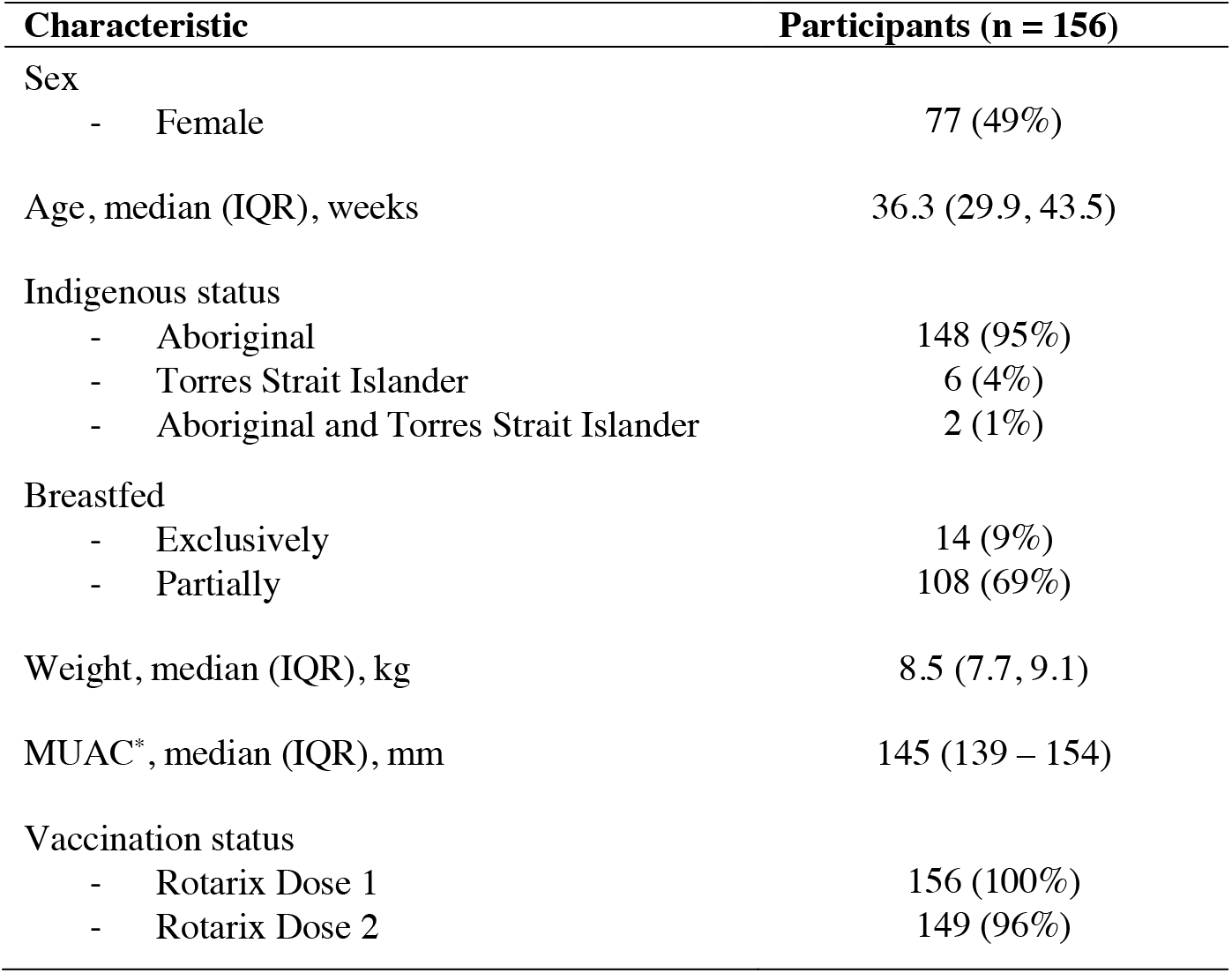
Baseline demographic characteristics including prior vaccine doses. *MUAC = mid-upper arm circumference

| Characteristic | Participants (n = 156) |
| --- | --- |
| Sex |  |
| - Female | 77 (49%) |
| Age, median (IQR), weeks | 36.3 (29.9, 43.5) |
| Indigenous status |  |
| - Aboriginal | 148 (95%) |
| - Torres Strait Islander | 6 (4%) |
| - Aboriginal and Torres Strait Islander | 2 (1%) |
| Breastfed |  |
| - Exclusively | 14 (9%) |
| - Partially | 108 (69%) |
| Weight, median (IQR), kg | 8.5 (7.7, 9.1) |
| MUAC*, median (IQR), mm | 145 (139 – 154) |
| Vaccination status |  |
| - Rotarix Dose 1 | 156 (100%) |
| - Rotarix Dose 2 | 149 (96%) |

Of the 156 participants, 119 (76%) were secretors and 129 (83%) were Lewis antigen positive (Table 2). Of the 8 samples that were indeterminate for secretor and Lewis antigen by phenotype assay, secretor status was identified by *FUT2* “G428A” mutation PCR assay for 7 samples. One sample was deemed ‘untypeable’.

**Table 2:** Rotavirus IgA seropositivity by Lewis and secretor status. ^*^Extra numeric indicates number of missing IgA values ^**^Geometric means are based on complete cases

| HBGA Phenotype | Participants<br>Proportion (%) | IgA Seropositive<br>≥ 20 AU/mL (n/%) | RR (95% CrI) | IgA GMC<br>AU/mL** |
| --- | --- | --- | --- | --- |
| **Secretor Phenotype** |  |  |  |  |
| Secretor | 119/156 (76%) | 87/119 (73%) (1)* | Reference | 68.8 |
| Weak Secretor | 9/156 (6%) | 4/9 (44%) | 0.62 (0.25, 1.02) | 42.6 |
| Non-Secretor | 27/156 (17%) | 13/27 (48%) (2)* | 0.66 (0.41, 0.93) | 24.6 |
| Untypeable | 1/156 (1%) | 1/1 (100%) | - | - |
| **Lewis Phenotype** |  |  |  |  |
| Positive | 129/156 (83%) | 89/129 (69%) | Reference | 56.2 |
| Negative | 19/156 (12%) | 10 (53%) | 0.79 (0.47, 1.10) | 56.4 |
| Untypeable | 8/156 (5%) | 6/8 (75%) | - | - |
| **Combined Phenotype** |  |  |  |  |
| Lewis Positive Secretors | 99/156 (63%) | 73/99 (74%) | - | 64.7 |
| Lewis Positive Weak Secretors | 9/156 (6%) | 4/9 (44%) | - | 52.6 |
| Lewis Positive Non-Secretors | 21/156 (13%) | 12/21 (67%) | - | 30.3 |
| Lewis Negative Secretors | 13/156 (8%) | 9/13 (69%) | - | 116.4 |
| Lewis Negative Non-Secretors | 6/156 (4%) | 1/6 (20%) | - | 9.2 |
| Untypeable/ Secretor | 7/156 (4%) | 5/7 (71%) | - | - |
| Untypeable | 1/156 (1%) | 1/1 (100%) | - | - |

Of the 156 participants, 105 (67%) were anti-rotavirus seropositive at baseline; 87 of 119 (73%) secretors versus 4/9 (44%) weak secretors and 13/27 (48%) non-secretors were seropositive, and 89 of 129 (69%) Lewis antigen positive versus 10 of 19 (53%) Lewis antigen negative were seropositive (Table 2). Crude (unadjusted) analyses, suggested that weak secretors and non-secretors were less likely to have evidence of IgA seropositivity compared to secretors, RR 0.62 (95%CrI, 0.25 – 1.02) and RR 0.66 (95%CrI, 0.41 – 0.93) respectively. Lewis negative participants were less likely to be seropositive than Lewis positive children, though evidence was weak RR 0.66 (95%CrI, 0.47 – 1.10)

## Discussion

The ORVAC clinical trial undertaken in Australian Aboriginal children provided an opportunity to examine for the first time the association between HBGA status and immune response to oral rotavirus vaccine in this high-risk population.

We found that a higher proportion of children with the secretor phenotype, compared with those with weak or non-secretor phenotype, were seropositive for anti-rotavirus antibodies following administration of their routine Rotarix vaccine schedule (73% vs. 44% and 48%, respectively). Higher rates of rotavirus vaccine IgA seroconversion and/or vaccine strain shedding (a marker of vaccine “take”) have been observed previously among secretors compared to non-secretors, in studies among children in Nicaragua, Pakistan, Ghana and Malawi.^4^ With the exception of a study in Nicaragua, which observed a lower proportion of seroconversion among Lewis-positive non-secretors after 1 dose of either Rotarix or RotaTeq,^4^ previous studies have not identified an effect of Lewis antigen status on vaccine response. We found no evidence of reduced seropositivity for this group, and instead observed reduced seropositivity for Lewis-negative non-secretors, though numbers were small (n=6).

While IgA seroconversion tends to broadly reflect oral rotavirus vaccine performance at the population level, individual level protection against rotavirus infection is mediated by both humoral and cellular components of the immune system.^9^ A study of 238 vaccinated children in Malawi found that the prevalence of non-secretor phenotype was significantly lower among children with vaccine failure (12%) compared to community controls with no diarrhoea (28%).^8^ The authors proposed that although non-secretor children might be resistant to live rotavirus vaccine strains, their resistance to naturally circulating wild-type rotaviruses might, on balance, nonetheless protect them from rotavirus disease. Similarly, a trial among children in Bangladesh observed that Rotarix provides similar protection to children with secretor and non-secretor phenotype, and that Lewis phenotype did not impact vaccine effectiveness.^10^

Recent studies have shown that HBGAs on the surface of enterocytes recognise the VP8* subunit of the VP4 outer capsid protein of the rotavirus particle, and may thus function as host receptors in the initiation of rotavirus infection; furthermore, this may occur in a genotype dependent manner.^4^ Globally, the rotavirus P-genotypes P[4], P[6] and P[8] predominate in children.^9^ The secretor phenotype has been associated with increased susceptibility to P[4] and P[8] rotavirus infections, while Lewis negative phenotype individuals have been shown to be preferentially infected by the P[6] genotype independent of secretor status.^4^ It is proposed that variation in the distribution of HBGA phenotype between populations and ethnic groups may account for some of the geographical variation in circulating rotavirus strains. For example, the high prevalence of Lewis negative phenotype in sub-Saharan Africa may explain why strains containing a VP4 P[6] are endemic there.^4^ Both oral Rotarix and RotaTeq rotavirus vaccines contain the VP4P[8] genotype and, considering the potential for HBGA-mediated, rotavirus genotype-dependent differences in rotavirus vaccine responses, it has been suggested that non-secretor children may respond poorly to these vaccines because of their intrinsic resistance to infection with P[8] rotaviruses.^4^ Furthermore, this may be one factor driving the different levels of vaccine efficacy seen in different populations.

Maternal HBGA status may play a role in rotavirus vaccine seroconversion among breast-feeding children. A recent study in Bangladesh reported rotavirus-specific IgA seroconversion rates were higher among breast-fed children of non-secretor mothers than among children whose mothers were secretors (39% vs 23% respectively).^11^ Although many children in our study were fully or partially breastfed at the time of enrolment, we are unable to ascertain maternal HBGA status or exposure to breastmilk at the time of prior oral rotavirus vaccine administration.

There were several limitations. In this cohort, most children were secretor positive (76%) and only a small proportion were Lewis negative (12%). While these data are consistent with the prevalence observed in European, North American and some Asian populations, and among non-Indigenous Australian children,^4,6^ they contrast with earlier studies of Australian Aboriginal adults where 97% of participants were reported to be secretor positive and the Lewis negative phenotype was reported to be as high as 58%.^12,13^ However, it is known that HBGA expression is developmentally regulated and so fucosyltransferase enzymes may not have yet reached normal activity levels in infancy,^14^ which may partly account for this discrepancy. In addition, some young children may express HBGA weakly. We used *FUT2* “G428A’ mutation PCR assays to confirm all indeterminate and non-secretor phenotypes identified by enzyme immunoassay. While the G428A is the most common *FUT2* mutation resulting in a non-secretor phenotype among Caucasians and Africans, a mutation at position 385 more commonly results in non-secretor phenotype among Asian people.^15^ We did not identify mutations other than at the 428 position in the *FUT2* gene and were unable to confirm Lewis status by sequencing the *FUT3* gene. Finally, the ORVAC study design meant that there was a delay between rotavirus vaccine administration (routine schedule 6 weeks and 4 months) and collection of serum samples for rotavirus antibody testing at age 6 to < 12 months old.

## Conclusion

Most Australian Aboriginal children were secretor and Lewis HBGA positive. Those with weak secretor and non-secretor phenotypes were less likely to be IgA seropositive after a routine schedule of oral rotavirus vaccine but the prevalence of these two phenotypes was low in this cohort. Thus, the influence of HBGA status is unlikely to fully explain the underperformance of rotavirus vaccine at a population level among Australian Aboriginal children.

## Data Availability

All data produced in the present study are available upon reasonable request to the authors

## Acknowledgments

The study investigators would like to acknowledge the valuable contribution of Aboriginal Elder, Ms Ada Parry, Aboriginal Members of the Steering Committee Dr Dennis Bonney, Dr Olivia O’Donoghue and Dr Simone Raye, and the Australian First Nations Reference Group for Child Health at Menzies school of Health Research and the Kalunga Aboriginal Research Development Unit at the Telethon Kids Institute for their contribution to the ORVAC study. We would also like to sincerely thank Dr Monica McNeal and her team at Cincinnati Children’s Hospital, USA for enabling training in the rotavirus IgA ELISA and their generous sharing of specialist reagents. This permitted the collected blood samples to remain on country in Australia, fulfilling the request from the local Child Health Indigenous Reference Group. Similarly, we would like to sincerely thank Prof Julie Bines and her team at the Murdoch Children’s Research Institute in Melbourne, for establishing the HBGA assay and processing the saliva samples within Australia, as requested by the local Child Health Indigenous Reference Group. Nigel Cunliffe is affiliated to the National Institute for Health and Care Research (NIHR) Health Protection Research Unit in Gastrointestinal Infections at the University of Liverpool, a partnership with the UK Health Security Agency in collaboration with the University of Warwick. Nigel Cunliffe is based at the University of Liverpool. The views expressed are those of the author(s) and not necessarily those of the NIHR, the Department of Health and Social Care or the UK Health Security Agency.

