## Supplemental Table 1 for "Histo-blood Group Antigen status of Australian Aboriginal children and seropositivity following oral rotavirus vaccination"

**Supplementary Materials**

**Table 1 Baseline demographic characteristics for participants in the ORVAC clinical trial (NCT02941107).**

| | **Rotarix (n= 128)** | **Placebo (n = 125)** |
|---|---|---|
| Sex | | |
| Male | 64 (50.0%) | 67 (53.6%) |
| Median Age (months; Q1-Q3) | 8.5 (6.9 – 10.3) | 8.7 (7.3 – 10.3) |
| Indigenous Status | | |
| Aboriginal | 123 (96.1%) | 121 (96.8%) |
| Torres Strait Islander | 5 (3.9%) | 2 (1.6%) |
| Aboriginal and Torres Strait Islander | 0 (0%) | 2 (1.6%) |
| Usual Location | | |
| Regional Urban | 31 (24.2%) | 33 (26.4%) |
| Remote | 97 (75.8%) | 92 (73.6%) |
| Breastfed | | |
| Exclusively | 10 (7.8%) | 8 (6.4%) |
| Partially | 89 (69.5%) | 97 (77.6%) |
| Not breastfed | 29 (22.7%) | 20 (16.0%) |
| Median Weight (kg; Q1-Q3) | 8.5 (7.7 – 9·4) | 8.5 (7.8 – 9.1) |
| Median Arm Circumference (mm; Q1-Q3) | 145 (140 – 155) | 144 (140 – 155) |
| Pre-enrolment Rotarix Doses | | |
| First dose | 128 (100%) | 125 (100%) |
| Median Age (weeks; Q1-Q3) | 6.6 (6.2 – 7.1) | 6.7 (6.3 – 7.6) |
| Second dose | 119 (93·0%) | 122 (97·6%) |
| Median Age (weeks; Q1-Q3) | 17.9 (17.3 – 18.8) | 17.7 (17.4 – 18.4) |
| Seroresponse at Baseline (IgA ≥ 20 AU/mL) | | |
| Yes | 83 (64.8%) | 92 (73.6%) |
| No | 37 (28.9%) | 29 (23.2%) |
| Missing | 8 (6.2%) | 4 (3.2%) |
| IgA Concentration at Baseline | | |
| Median (IU/mL; Q1-Q3) | 59.6 (17.8 – 151.0) | 93.1 (21.2 – 164.0) |
